# Pooling-in-a-pod: A Strategy for COVID-19 Testing to Facilitate Safe Return to School

**DOI:** 10.1101/2021.03.24.21254230

**Authors:** Ethan M. Berke, Lori M. Newman, Suzanna Jemsby, Bethany Hyde, Natasha Bhalla, Natalie E. Sheils, Nandini Oomman, John Reppas, Prateek Verma, Gerard A. Cangelosi

## Abstract

The COVID-19 pandemic prompted widespread primary and secondary school closures. Routine testing of asymptomatic students and staff, as part of a comprehensive program, can help schools open safely. “Pooling-in-a-pod” is a public health surveillance strategy whereby testing cohorts (pods) are based on social relationships and physical proximity. Pooled testing provides one laboratory result for the entire pod, rather than separate results for each individual. Pooling-in-a-pod allowed for weekly on-site point-of-care testing of all staff and students at an independent preschool to grade 12 school in Washington, D.C. Staff and older students self-collected anterior nares samples, and trained staff collected samples from younger students. Overall, 12,885 samples were tested in 1,737 pools for 863 students and 264 staff between November 30, 2020, and April 30, 2021. The average pool size was 7.4 people. Sample collection to pool result time averaged 40 minutes. Direct testing cost per person per week was $$24.24, including swabs. Four surveillance test pools were positive. During the study period, daily new cases in Washington, D.C., ranged from 10 to 46 per 100,000 population. A post-launch survey found most parents (90.3%), students (93.4%), and staff (98.8%) were willing to participate in pooled testing with confirmatory tests for positive pool members. The school reported a 73.4% decrease in virtual learning after program initiation. Pooling-in-a-pod is a feasible and cost-effective surveillance strategy that was acceptable to staff and families and may be appropriate for some schools. School officials and policymakers can leverage this strategy to facilitate safe, sustainable, in-person schooling.

## INTRODUCTION

The COVID-19 pandemic resulted in widespread closures of schools across the United States. Although these closures were intended to minimize the risk of disease transmission, studies have shown that they may have had an unintentional adverse impact on approximately 56.4 million school-aged children though reduced educational attainment and years of life lost.^1^ As of this writing, only about half of the student population is currently in the classroom, with the majority of those in hybrid learning models.^2^ However, some studies suggest that students in school may actually be at lower risk of COVID-19 exposure than students out of school either due to differences in transmissibility or through stricter enforcement of masking and physical distancing compared to home and community settings.^3^ Schools are now starting to plan for the 2021-2022 school year, and there are calls for a return to full in-person learning.^4, 5^ In addition to benefits for children from an academic and social standpoint,^6-9^ returning students to in-person learning carries considerable value for economically disadvantaged populations and women.^10, 11^ Schools will need to have strategies in place that allows the safe in-person attendance for students and staff and minimizes operational disruption.

Strategies to safely keep schools open include daily symptom screening, masking, physical distancing, extracurricular activity modifications, and optimization of facilities to minimize transmission.^12^ Vaccinations will also play a critical role in reduction and spread of disease. Unfortunately, it is unlikely that 100% of the staff and students will be fully vaccinated by the coming school year as a result of vaccine hesitancy and limited availability of vaccines for the youngest age groups. Therefore, asymptomatic spread, which may account for as much as 60% of transmission in the community and specific sub-populations, will continue to be a risk.^13^ An optimal re-opening strategy for schools should also include routine SARS-CoV-2 testing with a high-performing test for all students and staff with a turnaround time that allows for rapid and impactful decisions. Substantial challenges include access to testing, cost, turnaround time, and policies for addressing positive test results.^14^ Most schools do not currently have the resources or capacity to implement testing strategies for all.^15, 16^

Pooling of samples from multiple individuals is a strategy used by commercial or reference laboratories to increase efficiency^17^ If the pool yields a negative test result, all samples are assumed to be negative. If it is positive, additional testing is used to identify the infected individuals. By combining multiple samples in a single test, more people can be tested at a lower cost. Pooling is most cost-effective for low-prevalence diseases, where most pools are expected to be negative. Because sample dilution may reduce sensitivity, it is critical to use technologies with high analytical sensitivity.^18^

The traditional application of pooling generally does not consider pool composition on social or geographic relationships. In contrast, “pooling-in-a-pod” is a public health surveillance strategy in which cohort-specific testing pods are composed based on epidemiologic characteristics such as social relationships and physical proximity. Pooled testing provides one laboratory result for the pod, rather than separate results for each individual within the pod. In schools, pods may be classrooms or staffing clusters (e.g., cafeteria workers, administration team). Pooling-in-a-pod uses these natural relationships so that actions taken on a positive test result (e.g., contact tracing and confirmatory testing) can be similar for all pod members.

## PURPOSE

The goal of this demonstration was to evaluate the feasibility of a pooling-in-a-pod strategy to reduce the number of infections on campus, minimize testing resource requirements, and maintain continuity of operations, thereby enabling schools to safely operate in the COVID-19 era. The results will help to guide the development of site-appropriate testing strategies for COVID-19 as well as future infectious disease outbreaks.

## METHODS

The demonstration project was conducted in a not-for-profit, independent day school in Washington, D.C., with 904 children and 209 faculty and staff, plus additional contractors, on two campuses, operating in a hybrid learning model (1 week in person, 1 week off campus) for grade 1–12 students and fully in-person for preschool and kindergarten. Students had flexibility in moving between on-campus and distance learning. This project was conducted as an institutional review board-approved study with consent from parents and staff, and assent by students. The school and its research partners used intentional design principles to design the project, including outlining project leadership, goals, available resources, scenario planning, operations, and stakeholder engagement (Figure 1).

**FIGURE 1.**
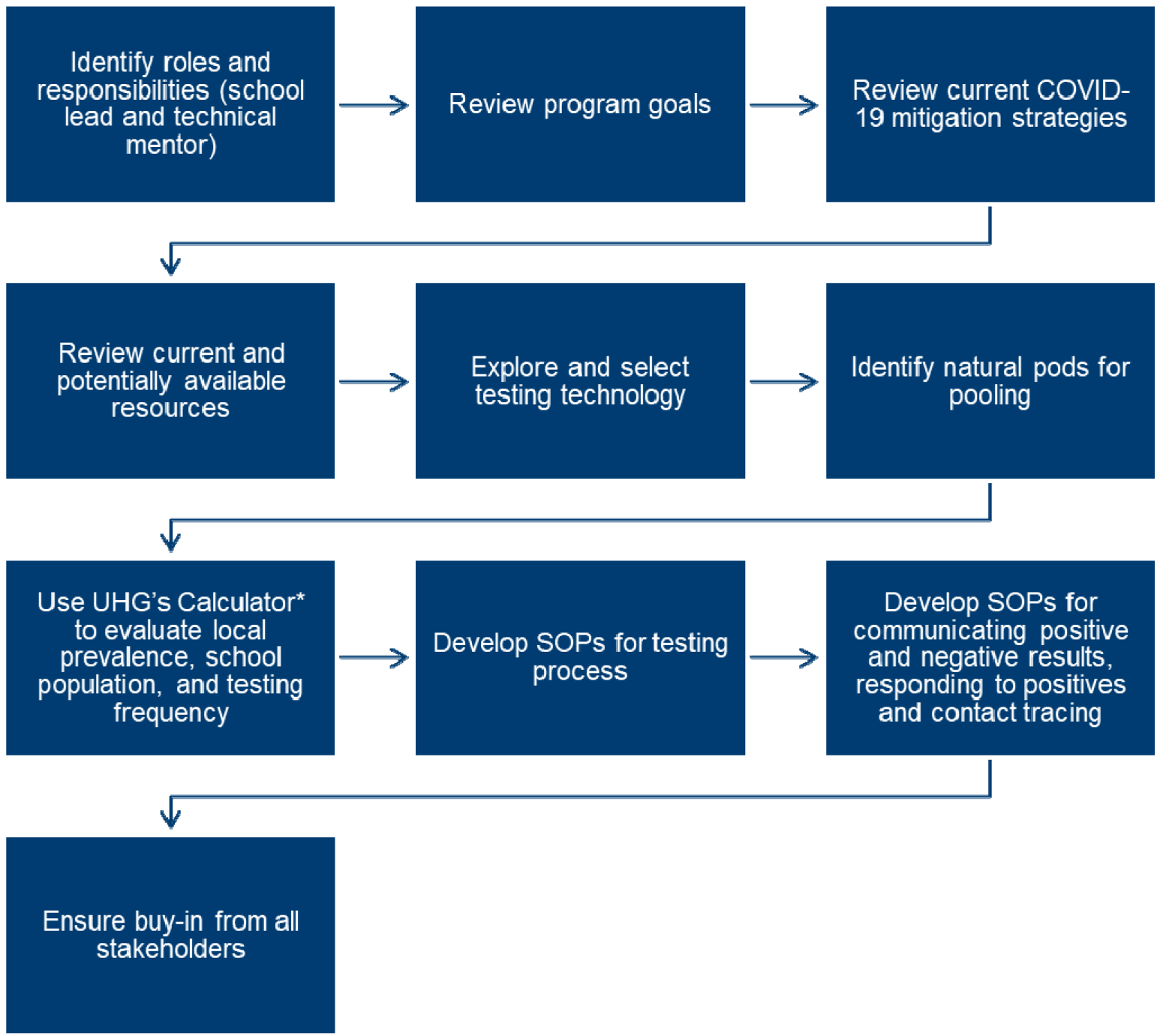
Design and implementation of a school-based pooling-in-a-pod strategy.

An online calculator^19^ was used to compare various hypothetical testing scenarios that allowed for the school to weigh the benefits of testing platform performance, testing frequency, cost, turnaround time, and pooling based on their operational constraints and program goals.^20^ Weekly testing plus symptom tracking with a $20, 60% positive agreement, 98% negative agreement test (e.g., individual antigen test in asymptomatic people) was estimated to cost $30.95 per person per week with confirmatory testing or $20.45 without and would reduce infections compared to symptom tracking only by 47% but result in 322 false positive results over 100 days. In contrast, a $175, 98% positive agreement, 99.5% negative agreement test (e.g., RT-PCR test) with same-day results administered weekly using pooling-in-a-pod with 14 people per pool was estimated to cost $21.57 per person per week with confirmatory testing, or $13.17 without, and would reduce infections by 98% but result in 82 false positive results over 100 days.

Based on this exercise, the school selected the portable, single-use Visby Medical COVID-19 test (Visby Medical, Palo Alto, CA), with performance similar to other nucleic acid amplification tests (NAATs),^21, 22^ but conducted on-site with a 30-minute turnaround time. The device was validated for pool sizes of 5 to 25; limit of detection was 2000 genomic copies/mL at 15 swabs per pool.^23^ The test was used according to federal guidance for pooled testing.^24, 25^ All swabs were introduced directly into a single buffer vial to minimize dilution during pooling.^18^ The target pool sizes of 8 to 14 for students and 4 to 6 for teachers/staff were based on class size, schedule, and estimated daily new cases in Washington, D.C. The range of 10 to 46 new cases per 100,000 population during the study period corresponded to a moderate to substantial community transmission risk.^26^ The school required persons to have a weekly negative test result to enter campus, either through the school pooled testing program at no cost to the participant, or through individual NAATs at the same frequency at their own expense. Alternatively, students could opt for distance learning only.

After a one-day, on-site training, the school operated all daily aspects including sample collection, device operation, data logging in secure software, and communications (Figure 2). Pods were designed at the school’s discretion to align to its operational needs and were composed of students only, staff only, or a mix depending on grade and schedule. Students tested twice per week when attending school in-person and did not test in their off week, resulting in an average testing frequency of once per week. Staff and younger students in full, on-site learning tested weekly. If a student or staff member was absent during their regular testing event, or if a substitute teacher came to campus mid-week, they were tested when they arrived on campus. Grades 6 to 12 students and all staff provided self-collected anterior nares samples^27^ and a trained clinician collected anterior nasal samples from students in preschool to grade 5.

**FIGURE 2.**
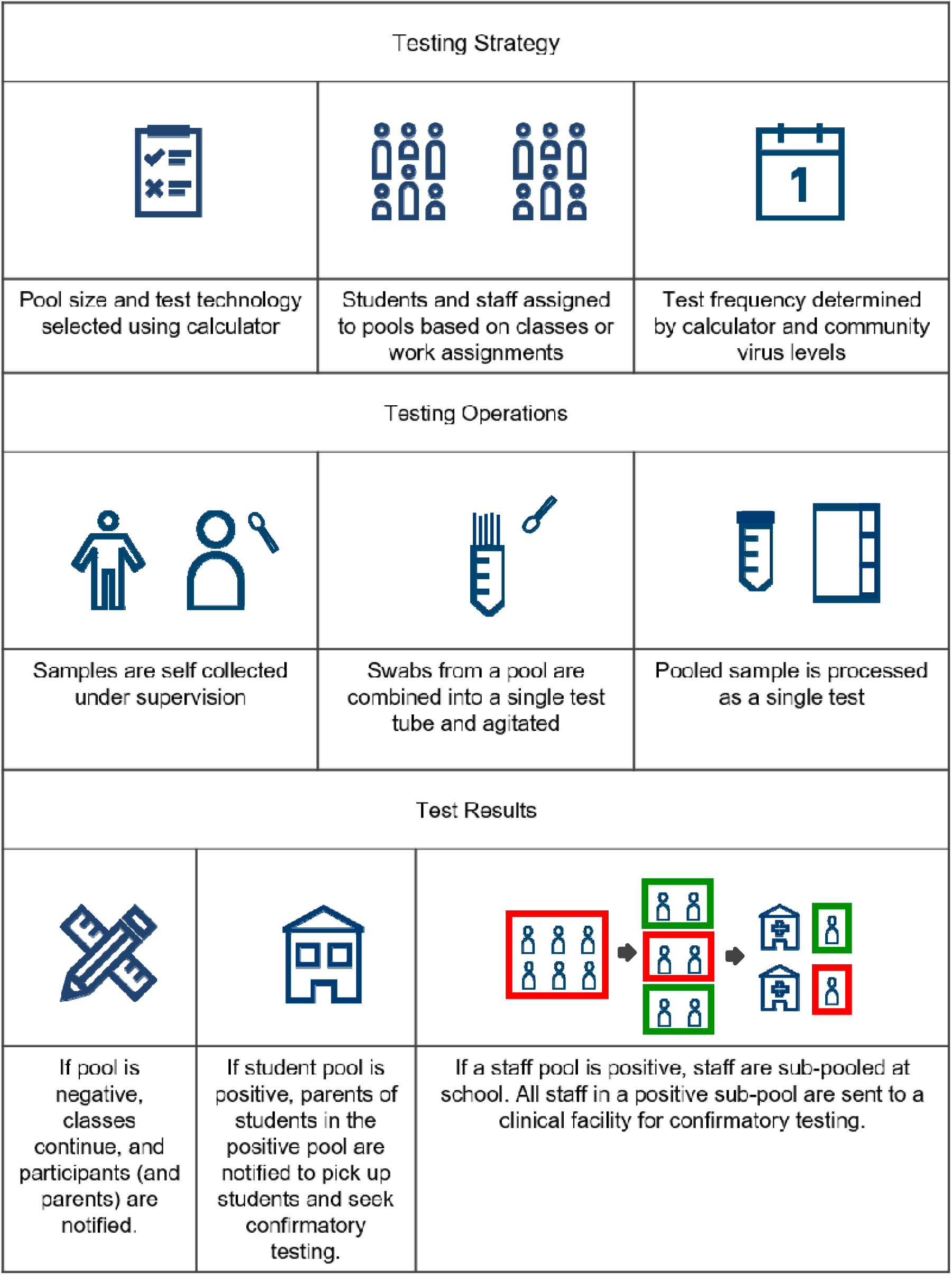
Operational flowchart for pooling-in-a-pod testing of faculty and students.

Pooled, not individual, results were communicated to staff and families via a single community-wide email update after each round of testing. If a pool was negative, all participants remained on campus. If a student pool was positive, students in that pool were sent home and advised to seek a NAAT in a clinical setting. If a staff pool was positive, all participants in that pool were asked to provide additional samples for sub-pooling, which minimized the number of staff adversely impacted by being in a positive pool. When a staff sub-pool was positive, members were confidentially advised to seek NAATs covered by employer insurance. Students and staff in a positive pool could not return until a negative NAAT result was available.

## OUTCOMES

From November 30, 2020, to April 30, 2021, 863 students and 264 staff and contractors participated at least once in the testing program (Table 2). Students in grades 6 to 12 started testing in November, while younger students did not participate until January 21, 2021. Grade 6 to12 students tested an average of 11.5 times (range 1 – 17), grade pre-K to 5 students tested an average of 10.6 times (range 1 – 17), and staff 12.7 times (range 1 – 25). Up to 542 students and 207 staff were tested each session; a total of 12,885 samples were tested in 1,737 pools. Of all students who came to campus during the study period, only one student chose to provide external proof of negative NAAT testing on a weekly basis instead of participating in the program. Average pool size was 7.4 (range 2 – 17). Testing time from sample collection to result averaged 40 minutes. Over 34 testing sessions in the study period, there were 1,733 negative and four positive pools. One pool of four staff was positive; outside individual confirmatory testing with NAAT identified a single positive person. A second positive pool of four staff was determined to be a false positive based on follow-up sub-pooling and outside confirmatory testing with NAAT. There were two additional positive pools of students; all students provided individual confirmatory NAAT results and in each positive pool a single asymptomatic positive student was identified. One individual reported a positive outside test during the study period, 4 days after a negative pooled test. Four people tested positive over the holiday break when no school testing was being performed. There were no confirmed cases of transmission within the school. The weekly direct per-person cost of the program was calculated identifying the cost per person per week, including swab, and applying a weighted average to calculate the overall per person cost. The weekly direct per-person cost of the program was $24.24.

**TABLE 1.** Parent, student, and staff attitudes to pooled testing in a survey administered three weeks after program initiation.

|  | AGREE or STRONGLY AGREE (%) |  |  |
| --- | --- | --- | --- |
|  | **PARENTS**<br>**n=309** | **STUDENTS**<br>**(Grades 6-12)**<br>**n=88** | **STAFF**<br>**n=84** |
| Response Rate (%) | 24 | 19 | 38 |
| QUESTION |  |  |  |
| Testing students, staff, and faculty on a regular basis is important to ensure that school can remain open and the WIS community can be as safe as possible. | 92 | 95 | 93 |
| Pre-launch: I am open to being part of a pooled testing protocol once or twice a week, with an individual confirmatory test required if the pool is positive | 89 | 88 | 92 |
| Post-launch: I am open to being part of a pooled testing protocol once or twice a week, with an individual confirmatory test required if the pool is positive | 90 | 93 | 99 |
| I feel that students or faculty who refuse to be tested individually or as part of a pool on a frequent basis should not be allowed to attend in person classes. | 80 | 83 | 74 |
| After being trained, I am comfortable collecting my own sample under observation at the school. | N/A<sup>1</sup> | 88 | 96 |
| I believe the testing program increases my comfort with the school moving toward full, in-person learning, even if other schools in the area remain in hybrid or stay-at-home models. | 82 | 76 | 65 |
| Now that I have been tested, I believe it is just as important to wear a mask, wash hands, and maintain physical distancing. | N/A | 92 | 96 |

<sup>1</sup> N/A: Not Applicable

**Table 2.** Student and staff pod attributes and pool results.

| **PODS** | **Number**<br>**Participating**<br>**at least once** | **Number of**<br>**Sessions**<br>**(mean, range)** | **Average Pool**<br>**Size (range)** | **Total Pools**<br>**(number**<br>**positive)** |
| --- | --- | --- | --- | --- |
| **STAFF** | 264 | 12.7 (1-25) | 6.62 (2-13) | 418 (2) |
| **STUDENTS**<br>**GR 6–12** | 442 | 11.5 (1-17) | 7.6 (2-17) | 670 (0) |
| **STUDENTS**<br>**PRESCHOOL–**<br>**GR 5** | 421 | 11.6 (1-17) | 8.1 (3-16) | 160 (2) |
| **MIXED**<br>**STUDENT &**<br>**STAFF** | N/A | N/A | 10.8 (2-17) | 489 (0) |
| **TOTAL** | 1127 | 11.8 (1-25) | 7.4 (2-17) | 1737 (4) |

Parents of grade 6 to 12 students and staff were surveyed after three weeks of testing; 309 parents, 88 students, and 84 staff responded (Table 1). After the program was launched, the majority of parents (90%), students (93%), and staff (99%) were open to participation. Parents, students, and staff reported increased comfort with in-person learning (82%, 76%, 65% respectively). Comments included the need for accurate, rapid results; a testing program that included everyone on campus; and minimized disruption to learning. Concerns centered on privacy, confidentiality, or responsibility for confirmatory testing. Prior to implementation on November 30, 2020, 90 of grade 6 to 12 students were in a distance learning model. As of April 30, 2021, only 34 remained in a distance learning model; similarly, as of November 30, 2020, 53 primary school students were in distance learning and as of April 30, 2021, four students were in distance learning. This corresponded to a 62.2% decrease in virtual learning for 6 to 12 grade students (p < 0.00001) and a 92.4% decrease in virtual learning for preschool to grade 5 students after initiation of the program (p < 0.00001).

## LESSONS LEARNED

Pooling-in-a-pod allows for more accessible COVID-19 testing in primary and secondary schools. This approach balances cost and convenience while optimizing turnaround time, frequency, and performance compared to other testing strategies such as non-pooled approaches. The program has a high acceptance and increases comfort with in-person attendance. It enables maximal on-campus learning within the framework of local restrictions. This program identified asymptomatic infection, possibly averting ongoing transmission.

Pooling-in-a-pod reduces costs and increases throughput. By assembling pods based on social networks and geography, particularly when coupled with rapid turnaround time, schools can make rapid decisions that can preserve continuity of operations. Pooled testing reduces the number of tests required, and therefore the cost of screening for asymptomatic positive cases in a school. Although this school required individual confirmatory diagnostic testing (and shifted this cost to insurance or publicly funded testing programs), other schools may instead use quarantine or isolation to further reduce organizational costs.^28^

It is widely believed that COVID-19 will become an endemic disease with intermittent regional outbreaks.^29^ Even as progressively larger numbers of teachers and students are vaccinated, vaccination of all school children will take time. Not all members of a school community may be vaccinated, and it is not yet clear what the risk of asymptomatic shedding is among vaccinated individuals, the role variants may play in asymptomatic transmission, and if booster vaccines will be required. Given the increasing body of evidence suggesting negative effects of remote learning on students, families, and society, and the expected presence of COVID-19 in the community, introduction of school surveillance testing programs may be an important investment to fully open school in the fall and stay open through the school year, complementing other mitigation efforts that include vaccinations.

This program was implemented with only one month of lead time. This could be shortened through adaptation of existing protocols and educational materials. Pooling-in-a-pod could potentially be scaled up rapidly with funding, leadership, and support from federal, private, and non-profit partners in health and education, even in settings such as public schools where implementation and workforce capacity are more limited. Rapid rollout of pooling-in-a-pod may help in the public health response to future pandemics as well. Despite positive acceptance of the program by the school community, achieving high participation rates to identify asymptomatic cases may require re-examination of school policies that mandate testing to be on campus, or utilizing an opt-out as opposed to opt-in consent processes.

This study had several limitations. The overall high approval of the program may not be generalizable to other settings and may be biased by a low survey response rate. While the majority of the parents are upper income, many of the parents and staff do not speak English as a first language. As such, communication routinely included slides with clear directions, non-verbal images, and was conducted by multilingual staff in English, French, and Spanish. Indirect costs were not included in actual cost estimates, the primary indirect cost being program staffing. Many schools will require additional human and financial resources to implement a testing program than were required for this demonstration project. However, pooled testing can reduce the cost of a testing program through efficiency gains. On-site or near-site high-throughput testing platforms may reduce costs further with a minimal loss of turnaround time. Tools that allow comparisons of cost, performance, and test frequency, such as online calculators,^19^ assist schools to make these strategic decisions.

Pooling-in-a-pod is a cost-effective, feasible, and acceptable surveillance testing strategy for primary and secondary schools to safely operate in-person when included as part of a comprehensive package of interventions to reduce transmission of SARS-CoV-2. Other innovations, including on-site and near-site dedicated labs, should be developed to facilitate national scale-up for all children. Pooling-in-a-pod public health surveillance could also be implemented for businesses and other institutions where in-person presence is essential.

## Data Availability

De-identified data are available for review.

https://calculator.unitedinresearch.com/

## ACKNOWLEDGEMENTS

Students, parents, and staff of the Washington International School; Bruce Tromberg, PhD, National Institute of Biomedical Imaging and Bioengineering (NIBIB), National Institutes of Health (NIH); RADM Michael Iademarco, USPHS and Centers for Disease Control and Prevention

